# Hydroxychloroquine and Tocilizumab Therapy in COVID-19 Patients – An Observational Study

**DOI:** 10.1101/2020.05.21.20109207

**Authors:** Andrew Ip, Donald A. Berry, Eric Hansen, Andre H. Goy, Andrew L Pecora, Brittany A Sinclaire, Urszula Bednarz, Michael Marafelias, Scott M. Berry, Nicholas S. Berry, Shivam Mathura, Ihor S Sawczuk, Noa Biran, Ronaldo C Go, Steven Sperber, Julia A Piwoz, Bindu Balani, Cristina Cicogna, Rani Sebti, Jerry Zuckerman, Keith M Rose, Lisa Tank, Laurie G Jacobs, Jason Korcak, Sarah L. Timmapuri, Joseph P. Underwood, Gregory Sugalski, Carol Barsky, Daniel W. Varga, Arif Asif, Joseph C Landolfi, Stuart L Goldberg

## Abstract

**Background:** Hydroxychloroquine has been touted as a COVID-19 treatment. Tocilizumab, an inhibitor of IL-6, has been proposed as a treatment of critically ill patients.

**Objective:** To describe the association between mortality and hydroxychloroquine or tocilizumab therapy among hospitalized COVID-19 patients.

**Design:** Retrospective observational cohort study of electronic health records Setting: 13-hospital network spanning the state of New Jersey.

**Participants:** Patients hospitalized between March 1, 2020 and April 22, 2020 with positive polymerase chain reaction results for SARS-CoV-2. Follow up was through May 5, 2020.

**Main Outcomes:** The primary outcome was death.

**Results:** Among 2512 hospitalized patients with COVID-19 there have been 547 deaths (22%), 1539 (61%) discharges and 426 (17%) remain hospitalized. 1914 (76%) received at least one dose of hydroxychloroquine and 1473 (59%) received hydroxychloroquine with azithromycin. After adjusting for imbalances via propensity modeling, compared to receiving neither drug, there were no significant differences in associated mortality for patients receiving any hydroxychloroquine during the hospitalization (HR, 0.99 [95% CI, 0.80-1.22]), hydroxychloroquine alone (HR, 1.02 [95% CI, 0.83-1.27]), or hydroxychloroquine with azithromycin (HR, 0.98 [95% CI, 0.75-1.28]). The 30-day unadjusted mortality for patients receiving hydroxychloroquine alone, azithromycin alone, the combination or neither drug was 25%, 20%, 18%, and 20%, respectively. Among 547 evaluable ICU patients, including 134 receiving tocilizumab in the ICU, an exploratory analysis found a trend towards an improved survival association with tocilizumab treatment (adjusted HR, 0.76 [95% CI, 0.57-1.00]), with 30 day unadjusted mortality with and without tocilizumab of 46% versus 56%.

**Conclusions:** This observational cohort study suggests hydroxychloroquine, either alone or in combination with azithromycin, was not associated with a survival benefit among hospitalized COVID-19 patients. Tocilizumab demonstrated a trend association towards reduced mortality among ICU patients. Our findings are limited to hospitalized patients and must be interpreted with caution while awaiting results of randomized trials.

**Trial Registration:** Clinicaltrials.gov Identifier: NCT04347993

## Introduction

The global pandemic caused by a novel coronavirus [severe acute respiratory syndrome (SARS)-CoV-2] and its disease, COVID-19, has led to infection in over 3 million individuals and more than 320,000 deaths as of May 21, 2020 [1]. In the United States there are currently over one million documented cases and more than 88,000 deaths [1,2]. As there are no approved treatments, management of COVID-19 is largely supportive [3,4].^4^

One empirical treatment for COVID-19 which has received attention is hydroxychloroquine, an antimalarial drug repurposed in recognition of its anti-inflammatory properties in the treatment of autoimmune conditions. Hydroxychloroquine and its analogue, chloroquine, demonstrate suppression of SARS-CoV-2 replication in vitro, with hydroxychloroquine demonstrating greater potency [5,6]. Studies from the original SARS-CoV virus suggest a mechanism of action involving impairment of the terminal glycosation of angiotensin converting enzyme 2 (ACE2), inhibition of SARS-CoV viral entry, and rapid elevation of endosomal pH that prevents endosome-mediated viral entry [7-10]. The immunomodulatory effects are thought to be due to the accumulation of the drug in lymphocytes and macrophages leading to reduction of proinflammatory cytokines, including type I interferons, tumor necrosis factor alpha, and interleukin-6 [9]. Other anti-inflammatory effects may be related to inhibition of signaling pathways [11].

Several small clinical reports have shown conflicting evidence regarding the efficacy of hydroxychloroquine in COVID-19 [12,13]. Recently an observational cohort study of 1376 hospitalized patients from a New York hospital using propensity modeling found no significant association between hydroxychloroquine use and intubation or death (hazard ratio, 1.04, 95% confidence interval, 0.82 to 1.32) [14]. A second observational cohort study of 1438 hospitalized patients throughout the New York metropolitan region also found a lack of survival association with hydroxychloroquine with or without concomitant azithromycin (HR 1.35 and 1.08 respectively) [15]. However, confirmation of these studies outside New York and the results of ongoing randomized clinical trials (RCT) are not yet available.

As the clinical course of COVID-19 progresses, patients enter a hyperinflammatory phase with dysregulation of adaptive immune responses and a cytokine storm with elevation in plasma levels of pro-inflammatory cytokines including interleukins (IL) 2,6, 7, and 10, granulocyte-colony stimulating factor (G-CSF), interferon-gamma-inducible protein-10 (IFN-gamma, IL-10), and tumor necrosis factor alpha (TNF-alpha). This cytokine storm results in a pro-thrombotic milieu, cardiomyopathy, and ultimately multi-organ failure [16,17]. Tocilizumab, a monoclonal antibody against membrane bound IL-6 receptor inhibiting binding of soluble IL-6 and subsequent signal transduction, has been proposed as a therapeutic candidate for impeding cytokine storm [18]. Small single institution series have suggested benefit among severely ill patients [19,20]. Preliminary results from a press release for the French CORIMUNO-TOCI trial (NCT04331808), an open-label randomized trial of hospitalized patients with COVID-19 (n = 129), noted a reduction in the proportion of participants who died or needed ventilation in the tocilizumab group, although full results of the trial have not yet been released [21].

In the absence of RCTs, observational studies may provide useful early insights into effective treatment strategies [22,23]. However in an observational study, treatment allocations are based upon physician judgement, rather than random assignment, increasing the risk of bias and not accounting for known and unknown risk factors. Thus, causal inferences on effectiveness of treatments are challenging, but confounding effects can be partially mitigated via statistical methods [24,25].

Understanding these limitations, but with the urgency for evaluating potential therapeutic approaches during the current COVID-19 pandemic, we established an observational database within a 13-hospital network spanning New Jersey using an integrated electronic health record (EHR) system (EPIC; Verona, WI). In this observational cohort study we report our survival outcomes with hydroxychloroquine and tocilizumab among hospitalized patients with COVID-19.

## Methods

### Study Design and Cohort Selection

This retrospective, observational, multicenter cohort within the Hackensack Meridian Health network (HMH) used EHR-derived data to study hospitalized COVID-19 patient outcomes. Our primary objective was to analyze the effect of hydroxychloroquine in hospitalized patients. A secondary, exploratory objective was to investigate the effect of tocilizumab in the ICU population.

Patients were included in the database based on the following inclusion and exclusion criteria: 1) Positive SARS-CoV-2 diagnosis by reverse-transcriptase polymerase chain reaction, 2) Hospitalized within the time frame of March 1, 2020 until May 5, 2020, 3) Non-pregnant, 4) Not on a randomized clinical trial, and 5) Did not die during first day of hospitalization, and 5) Were not discharged to home within 24 hours. (Fig. 1)

**Fig 1.**
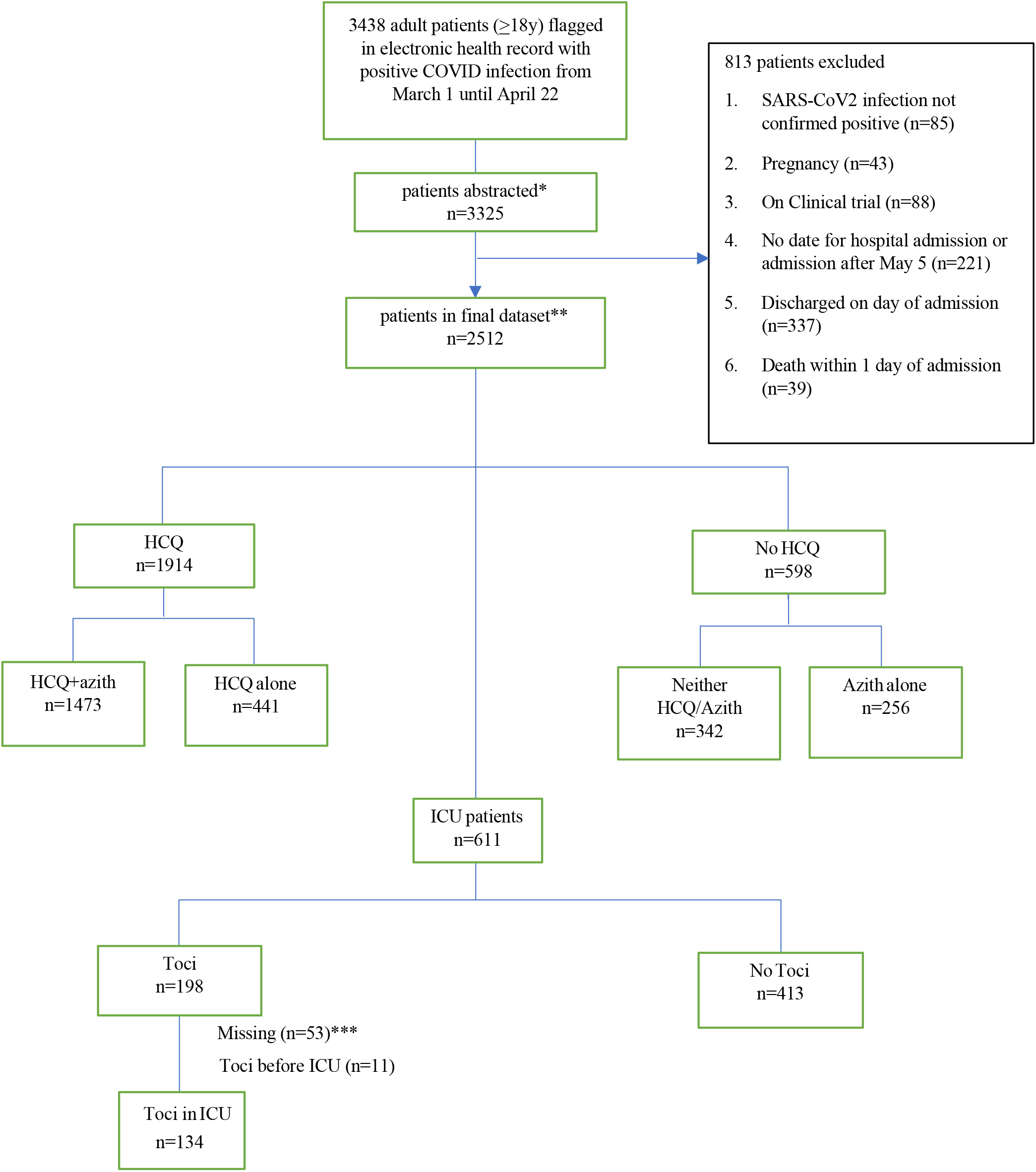
Cohort Selection Flow Diagram. Legend: Flow Diagram of patient sampling strategy of hospitalized COVID-19 patients in Hackensack Meridian Health network. *Convenience sampling was performed when assigning patients to our data team, and sampling bias is possible. 3325 of the 3438 (97%) available records were abstracted **Follow-up until final study cut-off date of May 5, 2020 *** 53 patients who received toci did not have sufficient data (date of administration) to analyze Tocilizumab (toci); Hydroxychloroquine (HCQ);

Institutional Review Board (IRB) approval was obtained for the observational database. Informed consent was waived by the IRB as this represented a non-interventional study of routinely collected data used for secondary research purposes. The study period was March 1, 2020 until May 5, 2020.

### Data Sources and Baseline Variables

We collected data from HMH’s EHR (Epic) which is utilized throughout the network. Hospitalized patients throughout HMH were flagged by the EHR if SARS-CoV-2 testing was positive. These EHR-generated reports served as our eligible cohort to sample. Demographic, clinical characteristics, treatments, and outcomes were manually abstracted by research nurses and physicians from the John Theurer Cancer Center at Hackensack University Medical Center. Assigning patients to our data team occurred in real-time, and not randomized. To reduce sampling bias, we abstracted 3325 of the 3438 (97%) possible hospitalized patients by April 22 (with follow up until May 5), and performed stratification as discussed in our analytic approach. Data abstracted by the team was entered using REDCap (Research Electronic Data Capture) hosted at HMH [26,27]. Data abstraction occurred daily from March 28 until May 5, 2020. Quality control was performed by physicians (AI, SLG) overseeing nurse or physician abstraction.

Demographic information was collected by an electronic facesheet, with gender, race or ethnicity self-reported. Academic centers were defined as quaternary referral centers with accredited residency, fellowship, and medical student programs. Nursing home or rehabilitation patients, if diagnosed prior to hospital admission, were defined as ambulatory patients. Comorbidities were defined as diagnosed prior to hospitalized for COVID-19. History of hypertension, diabetes, chronic lung disease (COPD or asthma), hypertension, cancer, coronary artery disease, cerebrovascular disease, renal failure, and rheumatologic disorder were abstracted from provider notes or medical history sections found within the EHR. If not listed in the patient’s record, the comorbidity was recorded as absent.

Presenting clinical data was abstracted from thorough review of unstructured notes as well as structured data. Hospital readmissions were counted as the same admission, with baseline data used from the initial hospitalization. If multiple positive or indeterminate results were found in a patient’s record for SARS-CoV-2, the first initial positive test was used as the date of diagnosis.

### Exposures

For the effect of hydroxychloroquine, we separated patients into 4 different groups – 1) Hydroxychloroquine, 2) Hydroxychloroquine in combination with Azithromycin, 3) Azithromycin alone, and 4) neither drug. Exposure to hydroxychloroquine or azithromycin was defined as documentation of drug administration in the EHR. Dosing, duration, and timing in relation to symptom onset and admission were also collected. If no evidence of administration of drug was found, this was recorded as not having received the drug.

For tocilizumab, exposure was defined as receipt of the drug within the ICU setting as found in the EHR. If no date of administration was found, this was labeled as insufficient missing data for analysis. If no evidence of administration of the drug was found, this was recorded as not having received the drug.

### Outcome Measures

The primary outcome measurement was death with follow-up through May 5, 2020. Mortality was identified on chart review by a provider note announcing time of death or if the EHR labeled the patient as deceased. As death certificates were not readily available, cause of death was identified using the EHR by identifying the most immediate cause(s) documented [28].

Respiratory cause of death included any hypoxic condition related to COVID-19. Cardiac cause of death included cardiac arrest, myocardial infarction, or arrhythmias. Infectious cause of death included bacterial sepsis or secondary infections not including COVID-19. Other cause of death included multi-organ failure as well as alternative causes. Follow-up occurred until the study cut-off date of May 5.

Adverse drug events related to hydroxychloroquine were also described, including discontinuation due to arrhythmia or QT prolongation. This was obtained by provider documentation as well as EKG reports within the EHR.

### Statistical Analyses

The statistical plan is available in the supplement. Descriptive analyses of baseline characteristics by hydroxychloroquine exposure were performed using chi-square tests for categorical variables. Dose, frequency, timing, and duration of treatment were also summarized.

We used propensity-score stratification for the remaining statistical analyses [29,30]. We fit a logistic regression model to the probability of being assigned to the experimental arm (tocilizumab, hydroxychloroquine, or hydroxychloroquine plus azithromycin) compared with the control population (not assigned to the respective treatment). Patients are stratified into propensity-score quintiles and these strata are used to adjust treatment effects in a proportional hazards model.

The model for selecting factors to be included in propensity scores was a two-stage backward selection approach. We evaluated each of the factors as univariate predictors with factors having p-value less than 0.10 included for further consideration. We removed factors sequentially and one at a time from the multivariate model if their p-values were less than 0.50, with largest p-values considered first. We fit the final propensity-scores model using multivariate logistic regression of the selected factors. We then stratified the propensity scores for the entire population into quintiles and used these quintiles as an ordinal (4-degree-of-freedom) variable to adjust the relative treatment comparisonin a proportional hazards model. (see Supplemental for output)

We evaluated the following factors for all propensity-score models: gender, coronary disease, stroke, heart failure, arrhythmia, African American, COPD,, renal failure, rheumatologic disorder, inflammatory bowel disease, advanced liver disease, age, diabetes mellitus, insulin use prior to hospitalization, asthma, HIV/hepatitis, any cancer, and log ferritin. The final propensity-score model for hydroxychloroquine included the first 15 of these factors. That for hydroxychloroquine plus azithromycin included the first 15 of these factors plus cancer. That for tocilizumab included age, gender, COPD, and renal failure.

Tocilizumab was assigned preferentially for patients in the ICU. Tocilizumab patients included in our analyses received their first dose of the drug in the ICU. Control patients were those who were admitted to the ICU and who never received tocilizumab either before or after admission to the ICU. The start time for analysis was the day of admission to the ICU. Hydroxychloroquine and hydroxychloroquine plus azithromycin were evaluated from day of hospital admission, whether initially in the ICU or not. The control population consisted of patients who never received the respective treatment.

We also sought to address the factorial nature of treatment with combination hydroxychloroquine and azithromycin. Propensity scores are based on predicting particular an individual therapy and so do not naturally generalize to a factorial setting. For these analyses we averaged the two propensity scores calculated separately for hydroxychloroquine and hydroxychloroquine plus azithromycin. We then stratified into propensity quintiles based on that average and proceeded as indicated above. The raw results of our proportional hazards analyses adjusting for propensity scores is provided. (Supplemental)

Patients still alive and in the hospital were censored as of May 5, 2020. Patients who had been discharged from the hospital were censored as of day 36 following hospital admission.

We provide hazard ratios of treatment in comparison with control together with corresponding p-values and confidence intervals based on Wald tests. We show the unadjusted survival data using Kaplan-Meier plots from which we identify 30-day mortality for each treatment. Statistical calculations used JMP® Pro 15.0.0.

Confidence intervals and p-values in this study are descriptive measures of distance between outcomes of treatment groups or distance from hazard ratio 1.00. These measures do not have the same inferential interpretations that are possible for primary end point analyses of RCTs.

## Results

### Unadjusted Baseline characteristics of hospitalized patients

There were 3,438 patients flagged in our EHR with positive COVID19 infection within the 13-hosptial network spanning New Jersey, and data was abstracted on 3,325 (97%). 2512 hospitalized patients met inclusion criteria for this study. (Figure 1) As indicated in Table 1, the median age of the cohort was 64 years (IQR 52-76) with a male predominance (62%). Nursing home and rehabilitation patients comprised 16% of the cohort. Co-morbidities were common with 55% having hypertension, 41% obesity (BMI ≥30), 32% diabetes, 16% coronary arterial disease, 15% COPD/asthma, 12% cancer, and 31% having 3 or more chronic conditions.

**Table 1.**
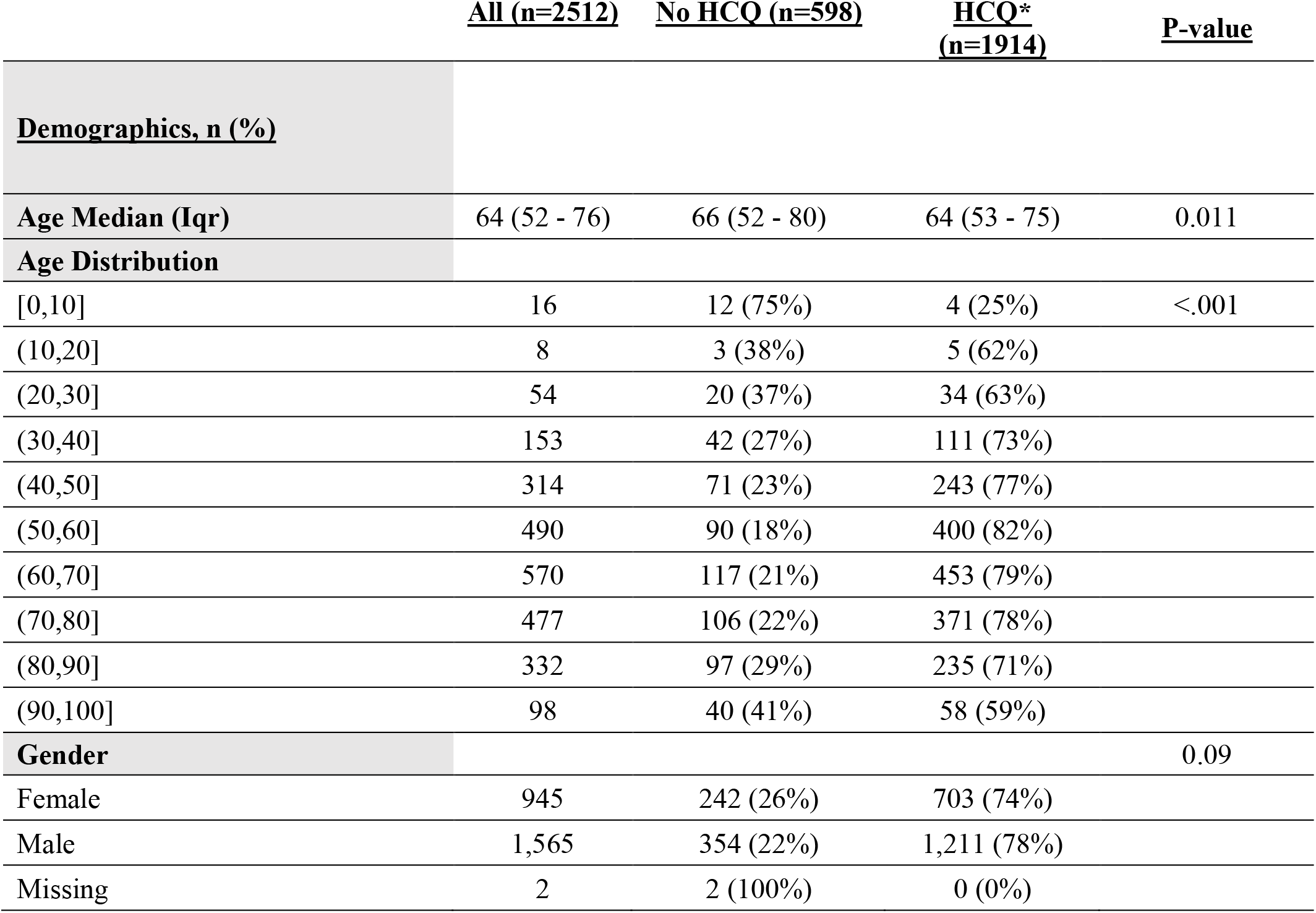
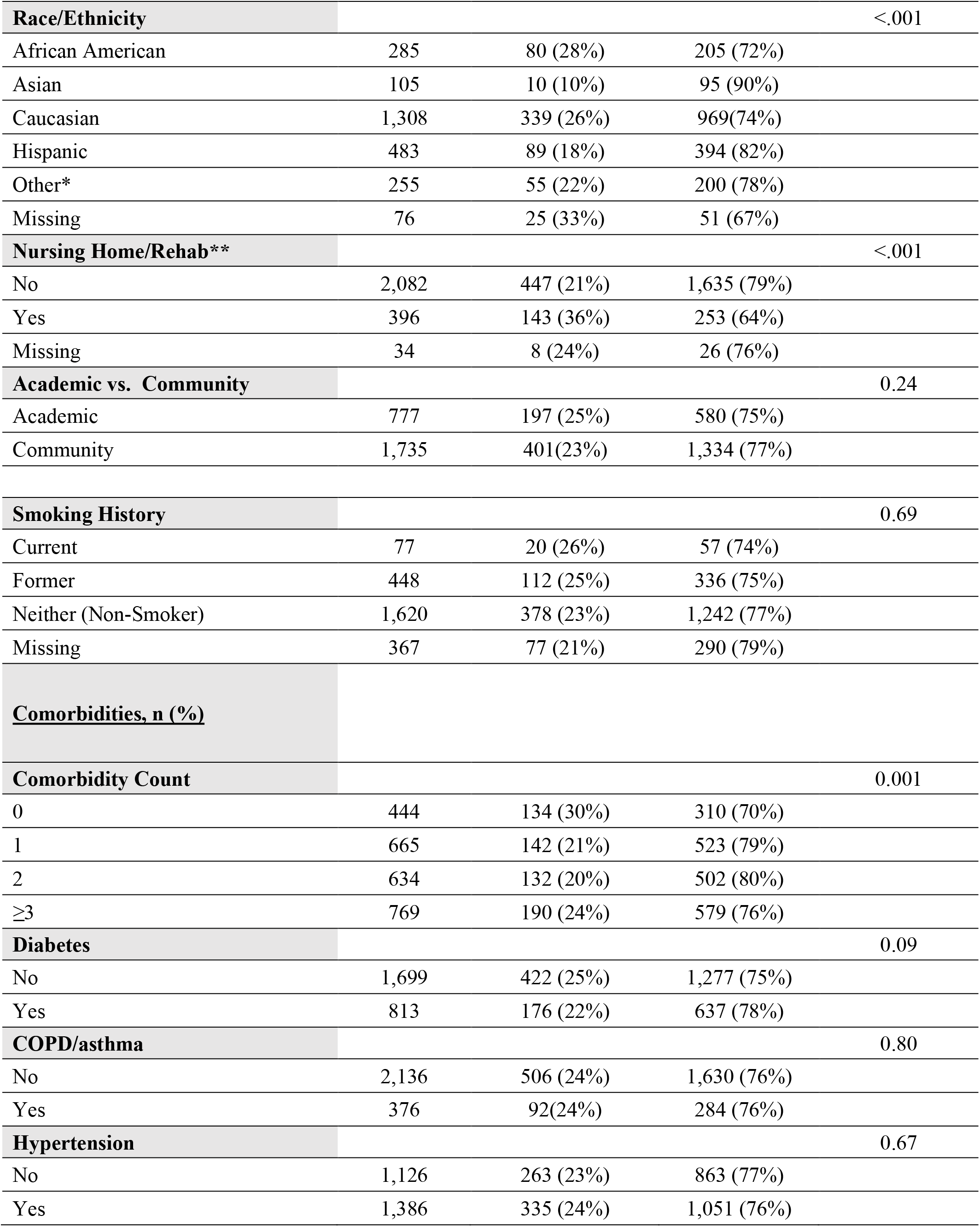
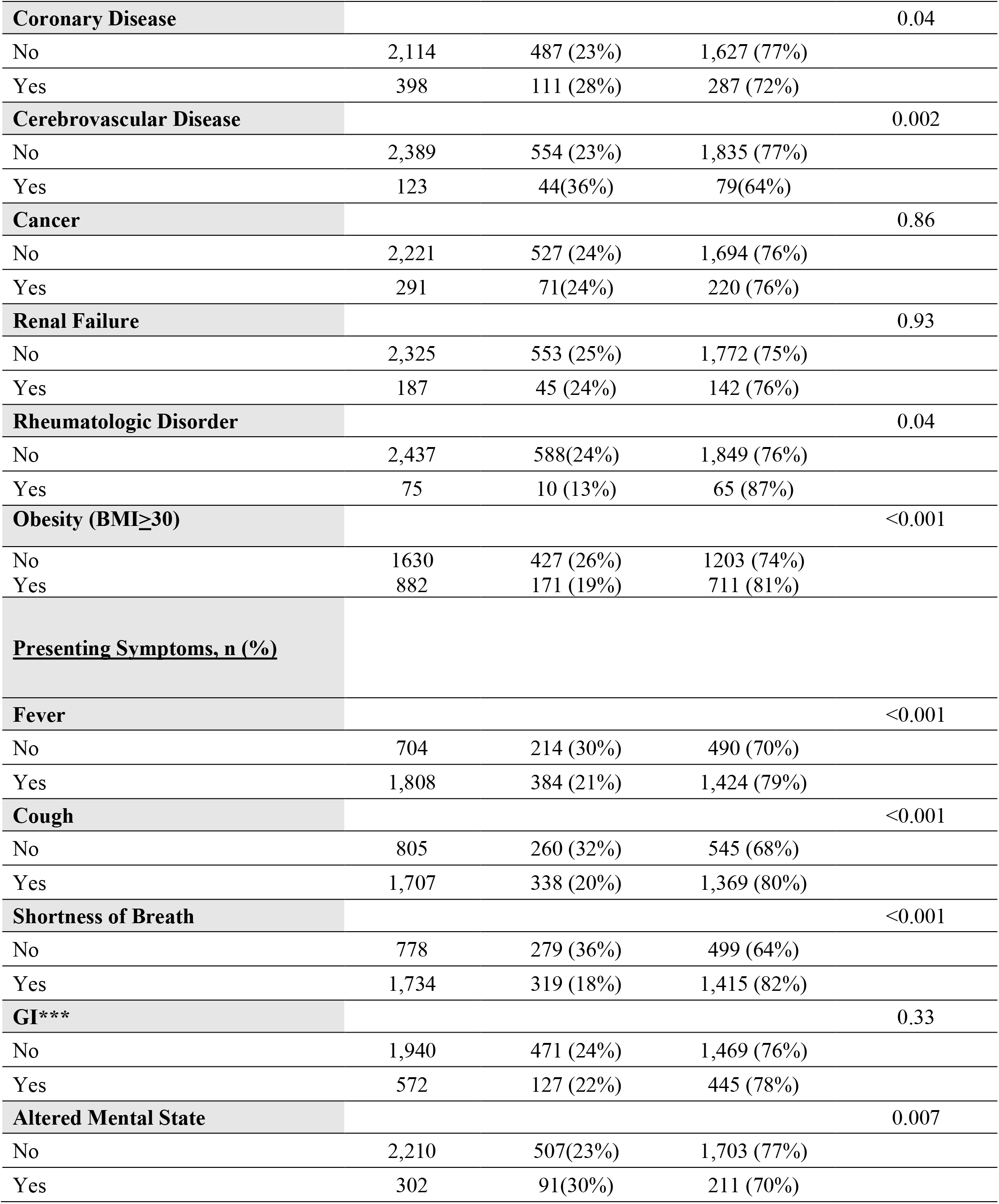
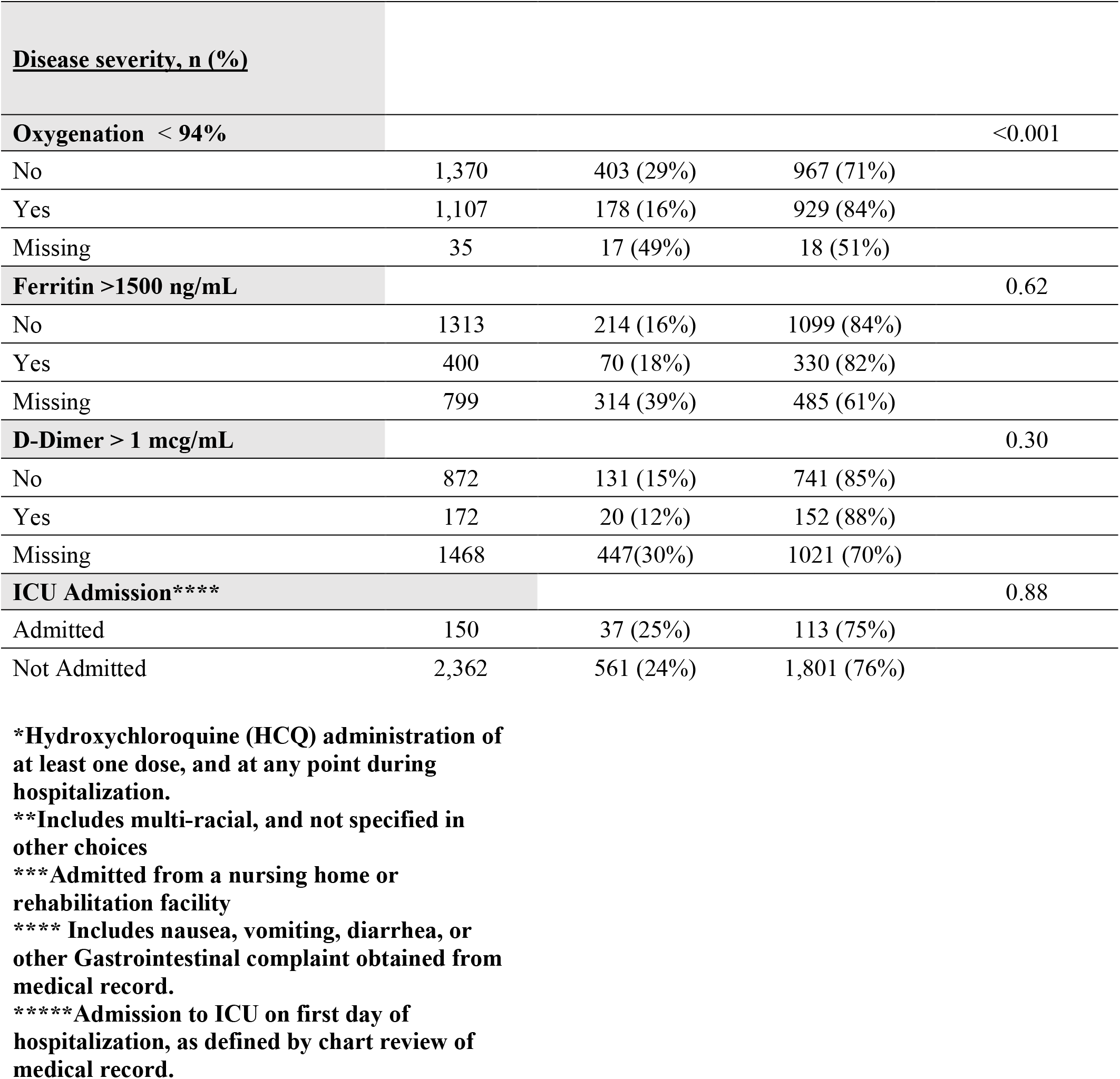
Unadjusted Baseline Characteristics of Hospitalized COVID-19 patients.

At the time of hospital presentation, fever was present in 72%, 69% with dyspnea, 68% with cough, 23% gastrointestinal complaints and 12% had altered mental status. The median time from self-reported onset of symptoms to hospitalization was 5 days (IQR 3-7). 611 (24%) patients required intensive care unit support during their hospitalization, of which 150 patients were admitted to the ICU within the first day. Oxygen saturation below 94% was identified in 44%. When measured and recorded in the electronic health record, inflammatory markers were elevated with serum ferritin >1500 ng/mL in 23% and d-dimer >1 mcg/mL in 16% of patients.

### Treatment with hydroxychloroquine

In this non-randomized observational cohort 1914 hospitalized patients (76%) received at least one dose of hydroxychloroquine during the study timeframe (23% as single agent and 77% in combination with azithromycin). (Figure 1) Dosing and duration was at prescribers’ discretion. The majority of patients received 800 mg on day 1, and 400 mg on day 2-5 (80%, n=1533), followed by 200 mg TID (4%, n=71) and other (15%, n =299), and missing dosing information (1%, n=11). Median duration of hydroxychloroquine was 5 days (IQR 4-5). The median days of symptoms before hydroxychloroquine administration was 5 (IQR 2-9), and median days in hospital before first dose 1 was 1 (IQR 0-2).

Patients receiving hydroxychloroquine at anytime during their hospitalization were younger, less likely to live in a nursing home, but presented later in their clinical course (5 days vs 3 days of symptoms prior to hospital admission) with more symptomatic disease (higher incidences of fever, cough, dyspnea, and lower oxygen saturation). (Table 1) After the FDA Emergency Use Authorization on March 28, 2020, prescriptions of hydroxychloroquine in our cohort temporally increased [569 (30%) prior to March 28 compared to 1314 (68%) post announcement, 42 (2%) dates missing] [31].

Discontinuation of hydroxychloroquine due to prolongation of QTc or arrhythmias was recorded in the electronic health records in 76 (4%) and 33 (2%) patients. During the entire hospital course, arrhythmias were noted in 101 (5%) hydroxychloroquine patients, compared to 22 (4%) in the non-hydroxychloroquine cohort. Cardiomyopathy was described in 20 (1%) treated patients compared to 7 (1%) patients without hydroxychloroquine.

### Survival Outcomes and Hydroxychloroquine Therapy

Among 2512 hospitalized patients with COVID-19 there have been 547 deaths (22%), 1539 (61%) discharges and 426 (17%) remain hospitalized as of the study cut-off date. In the entire cohort, causes of death included 319 (58%) respiratory, 108 (20%) cardiac, 42 (8%) other, 41 (7%) infectious, and 37 (7%) other. Within the hydroxychloroquine treated cohort, 89 of 432 (21%) of deaths were attributed to cardiac causes, compared to 19 of 115 (16%) deaths in the non-hydroxychloroquine cohort.

As shown in table 2, using propensity modeling as described above, there was no significant association between survival and any use of hydroxychloroquine during the hospitalization (HR, 0.99 [95% CI, 0.80-1.22]), hydroxychloroquine alone (HR, 1.02 [95% CI, 0.83-1.27]), or hydroxychloroquine in combination with azithromycin (HR, 0.98 [95% CI, 0.75-1.28]). The unadjusted 30-day mortality for patients receiving hydroxychloroquine alone, azithromycin alone, the combination or neither drug was 25%, 20%, 18%, and 20%, respectively. (Fig. 2)

**Table 2:** Propensity-score adjusted hazard ratios, confidence intervals

|  | HR | 95% conf. interval |  | P-value | 30-day mortality rate |  |
| --- | --- | --- | --- | --- | --- | --- |
|  |  | Lower | Upper |  | Experimental | Control |
| Any HCQ in hospital | 0.99 | 0.80 | 1.22 | 0.92 | 0.20 | 0.20 |
| HCQ+AZI in hospital | 0.98 | 0.75 | 1.28 | 0.89 | 0.18 | 0.20 |
| Factorial main effects and interaction of HCQ and AZI |  |  |  |  |  |  |
| HCQ main effect | 1.02 | 0.83 | 1.27 | 0.83 | 0.25<br>(HCQ only) | 0.20<br>(Neither) |
| AZI main effect | 0.89 | 0.72 | 1.10 | 0.28 | 0.20<br>(AZI only) | 0.20<br>(Neither) |
| Interaction |  |  |  | 0.091 | 0.18<br>(Both) | 0.20<br>(Neither) |
| Tocilizumab therapy in the Intensive Care Unit |  |  |  |  |  |  |
| Toci in ICU | 0.76 | 0.57 | 1.00 | 0.053 | 0.46 | 0.56 |

**Fig. 2:**
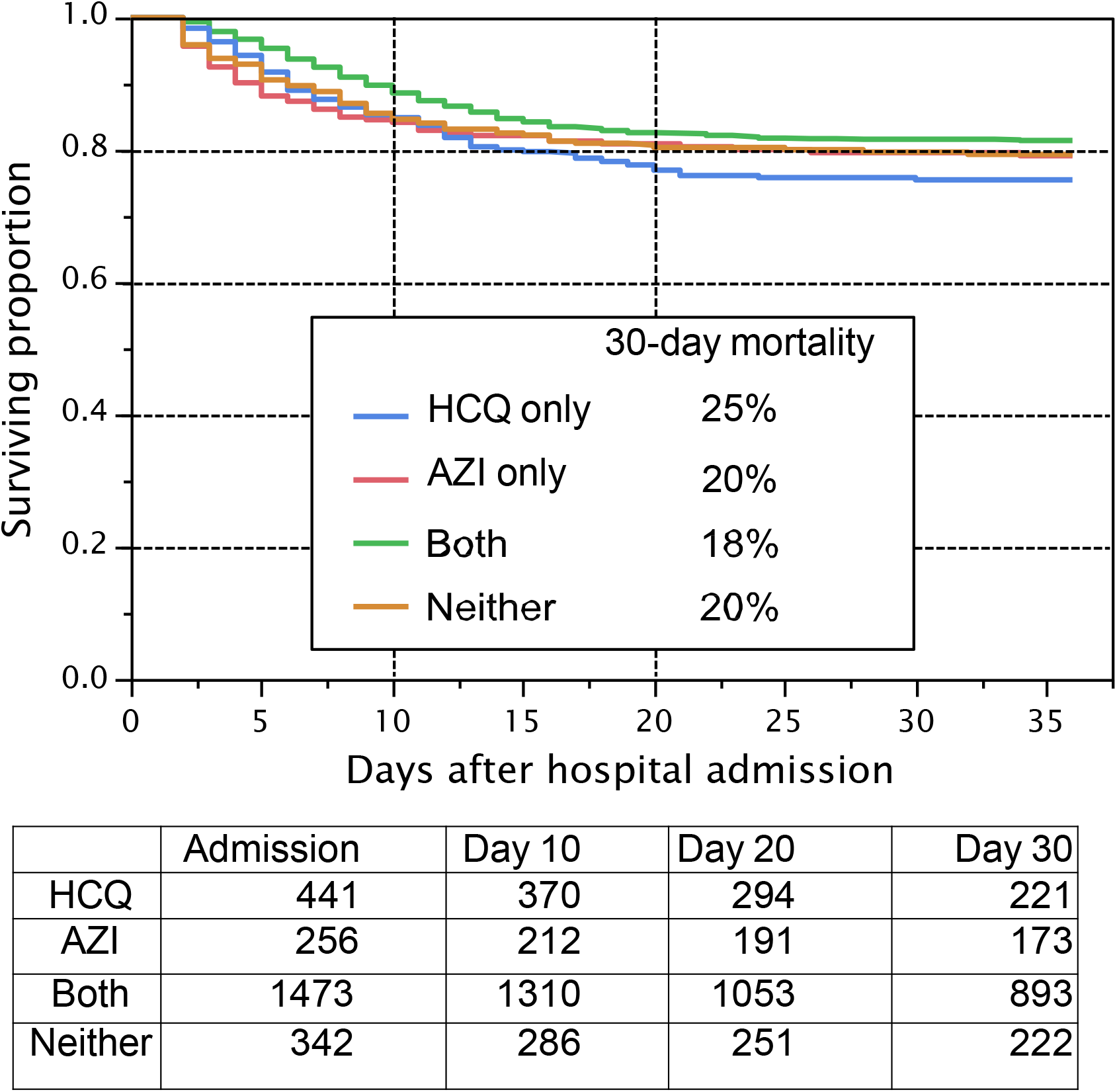
Unadjusted Association of Treatment with Hydroxychloroquine on Overall Survival Among Hospitalized COVID-19 Patients **Legend:** Unadjusted Kaplan-Meier estimates of survival by treatment allocation to hydroxychloroquine (HCQ), azithromycin (AZI), both, or neither. Patients at risk at admission, day 10, day 20, and day 30 are shown. Patients still alive and in the hospital were censored as of May 5, 2020. Patients who had been discharged from the hospital were censored as of day 36 following hospital admission. 30-day mortality rates are shown for each treatment.

### Exploratory Analysis on Tocilizumab Therapy

198 (8%) patients received tocilizumab therapy during their hospitalization. Table 3 shows baseline characteristics. 11 patients received their first dose prior to admission to the ICU and 53 patients had unknown timing or insufficient data. Thus 134 patients with documentation of their first dose of tocilizumab within an ICU setting represented the exploratory treatment cohort. 413 patients in the ICU never received tocilizumab as of May 5, 2020 and serve as the control cohort. Tocilizumab was administered as a single dose in 104 (78%), with the majority receiving 400 mg (96%), followed by 800 mg (1%), 8 mg/kg (1%), 4 mg/kg (1%), and missing dosing (1%). Secondary bacteremia occurred in 44 of the 413 (11%) patients in the non-treated group, compared to 18/134 (13%) in the treated group. Secondary pneumonia occurred in 25 of the 413 (6%) patients in the non-treated group, compared to 12 of the 134 (9%) in the treated group. As shown in table 2, in the analysis using propensity modeling, there was a trend association between survival and treatment with tocilizumab within the ICU setting (HR, 0.76 [95% CI, 0.57-1.00]). The unadjusted 30-day mortality favored tocilizumab (46% versus 56%). (Fig. 3)

**Table 3:**
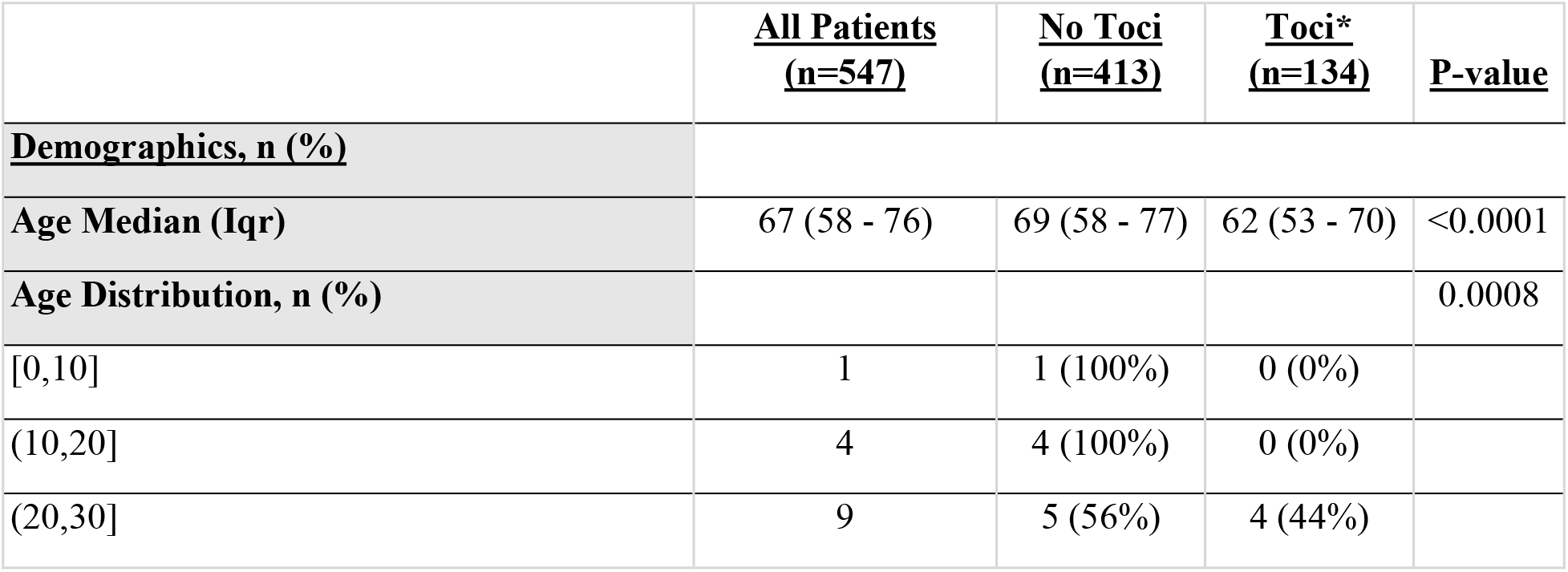
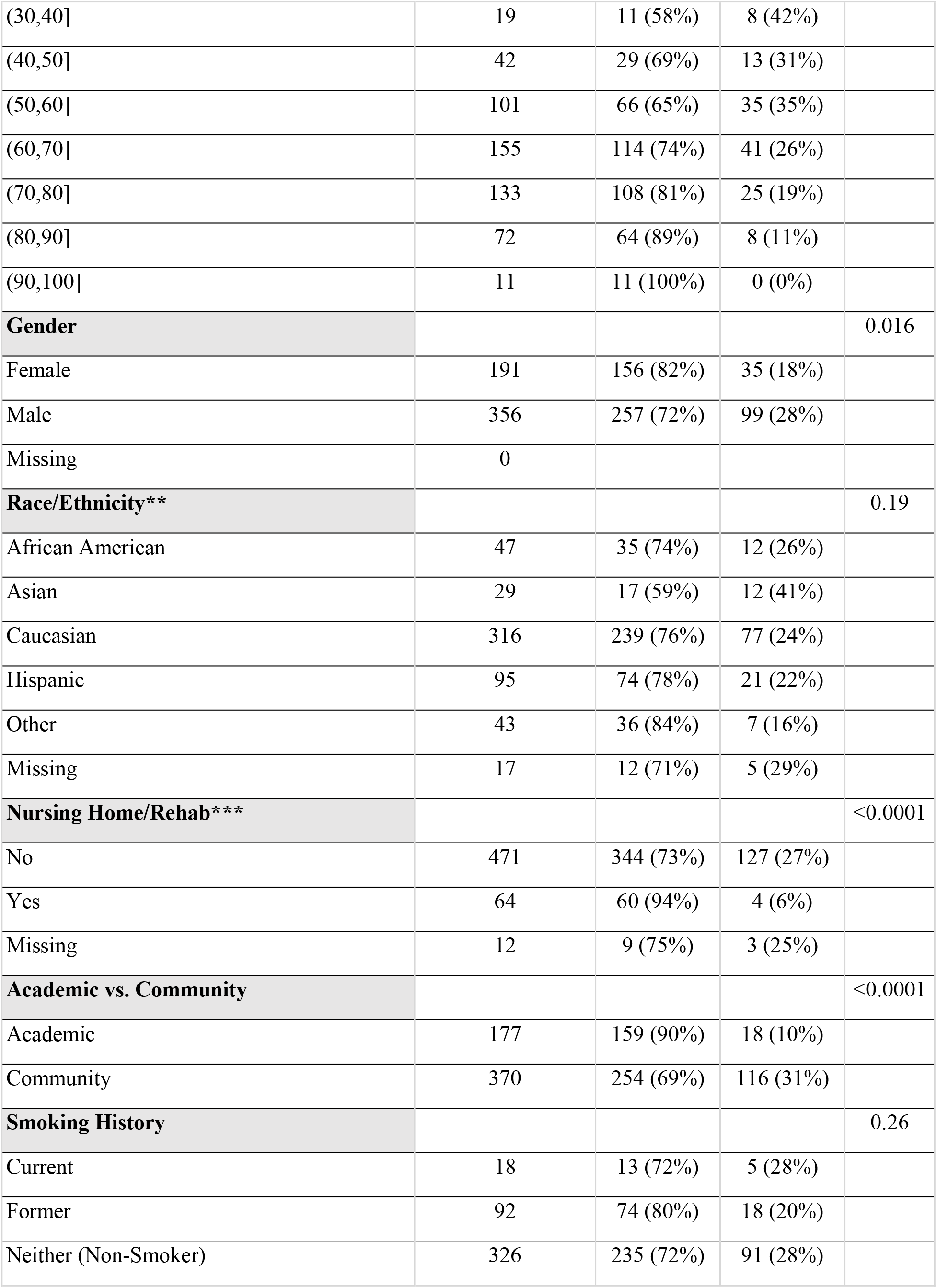
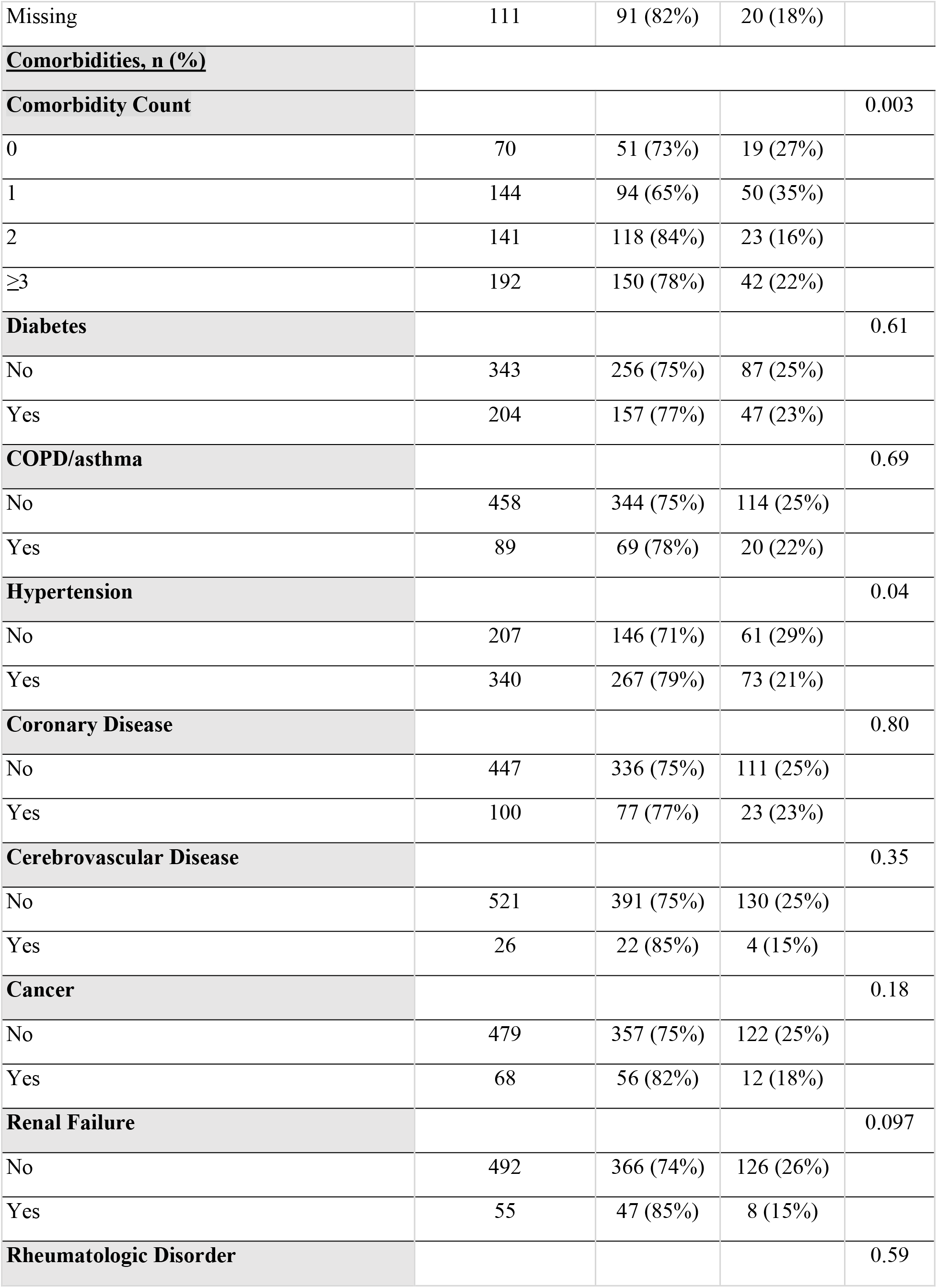
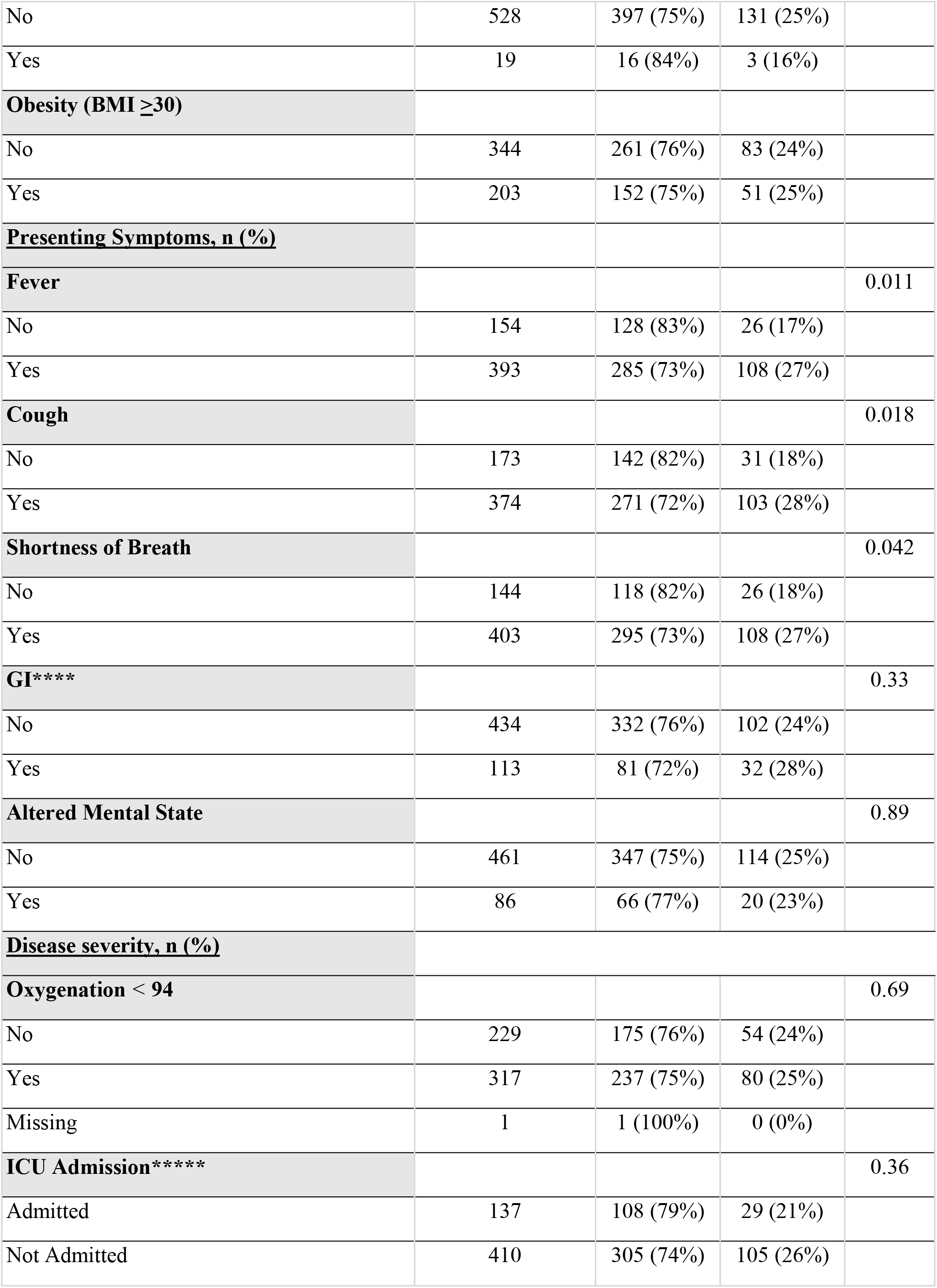
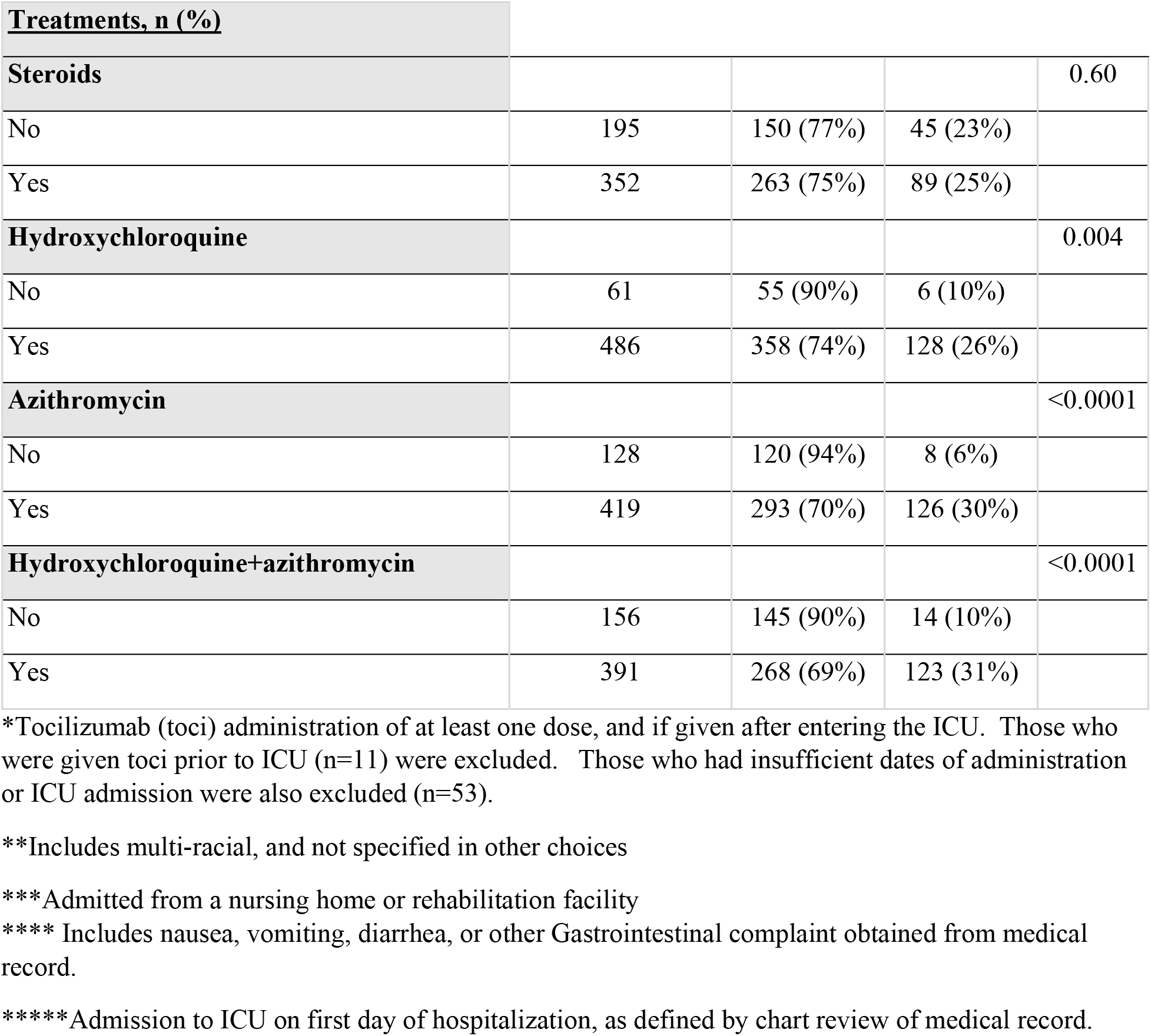
Unadjusted Baseline Characteristics of evaluable ICU patients receiving Tocilizumab.

**Figure 3:**
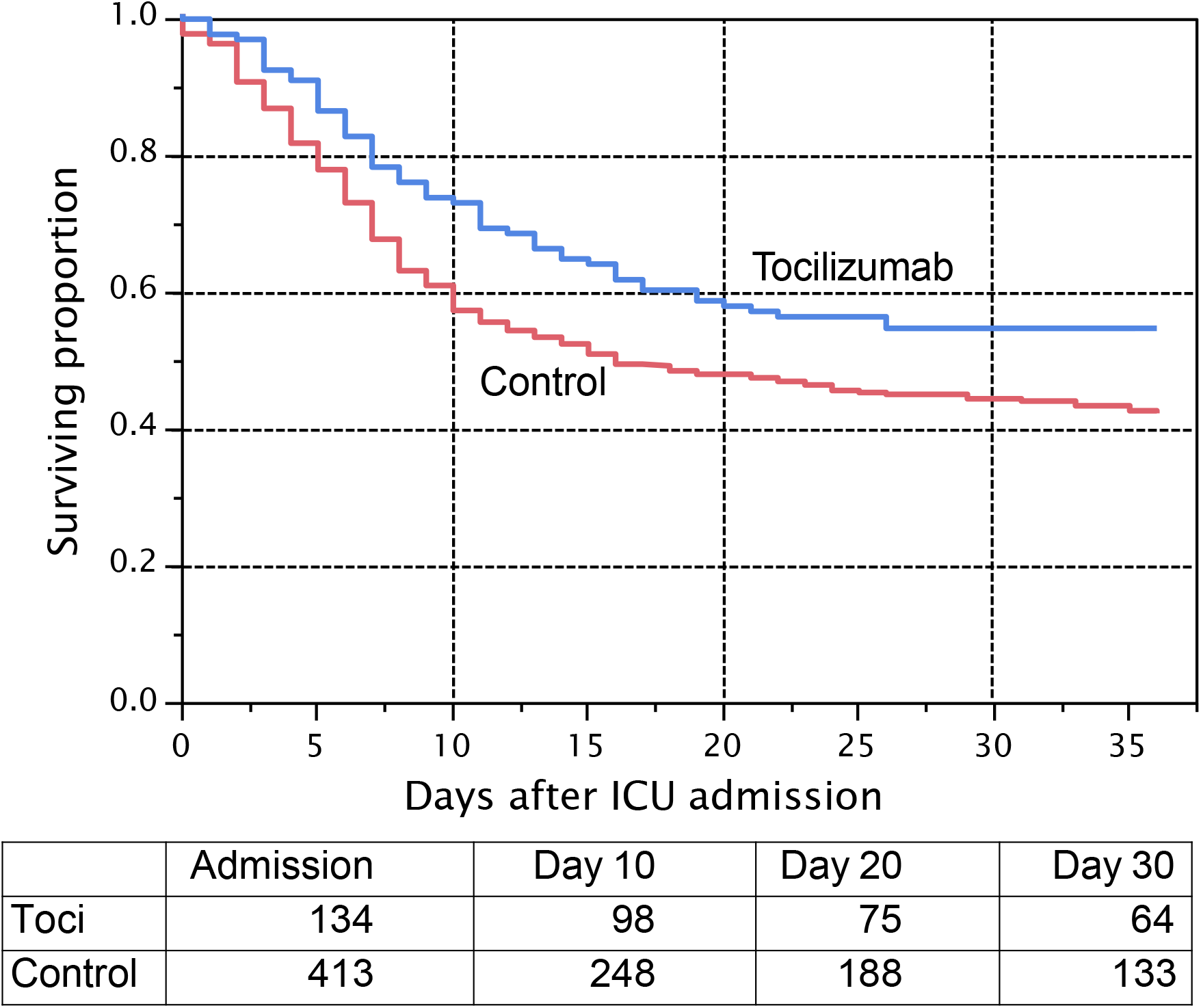
Unadjusted Association of Treatment with Tocilizumab on Overall Survival Among Hospitalized ICU COVID-19 Patients **Legend**: Unadjusted Kaplan-Meier estimates of survival by treatment allocation to tocilizumab (toci) or no toci. Patients at risk at admission, day 10, day 20, and day 30 are shown. Patients still alive and in the hospital were censored as of May 5, 2020. Patients who had been discharged from the hospital were censored as of day 36 following hospital admission. 30-day unadjusted mortality with and without tocilizumab is 46% versus 56%.

## Discussion

This retrospective observational cohort study of 2512 hospitalized COVID-19 patients within a 13-hospital network did not find the empirical use of hydroxychloroquine with or without cotreatment with azithromycin to be associated with a reduction in mortality (adjusted HR, 0.99 for any hydroxychloroquine during hospitalization [95% CI, 0.80-1.22]). Although our multi-center findings confirm the recent observational studies from New York, results of a randomized controlled study, the gold standard in determining drug efficacy, remain unavailable [14,15]. Nonetheless, collectively these observational studies do not support the routine use of hydroxychloroquine outside a clinical trial. Furthermore, none of the reported observational studies have addressed the role of hydroxychloroquine among individuals with minimally symptomatic disease in the pre-hospital setting.

Unlike randomized controlled trials, observational studies have inherent biases in patient allocations that cannot be fully adjusted for during statistical analyses. Although we utilized propensity modeling to mitigate known imbalances it is possible that unmeasured confounding factors may still be important. For example, in our series we observed a change in the prescribing patterns of hydroxychloroquine during the study timeframe. Similarly dosing and timing of hydroxychloroquine varied throughout the 13-hospital network. These factors (dosing, schedules, and changes in treatment patterns over time) are difficult to quantify in large observational studies. Nonetheless, our primary analysis is concordant with the other recent observational studies.

A potentially clinically important finding from an exploratory analysis revealed a favorable association between tocilizumab administration and survival among COVID-19 patients requiring ICU support. Our study represents the largest report of tocilizumab during this pandemic but is subject to all of the concerns of observational studies. However, if confirmed by ongoing randomized clinical trials, tocilizumab would represent the first successful therapy to reduce mortality.

### Limitations

Multiple RCT’s will ultimately determine the efficacy of hydroxychloroquine and tocilizumab in COVID-19. This observational study has limitations. First, observational studies cannot draw causal inferences given inherent known and unknown confounders. We attempted to adjust for known confounders using our propensity model approach. Second, misclassification is possible as we performed manual abstraction of EHR data. Third, low sample size limited our exploratory tocilizumab analysis. Fourth, our study focused on patients in New Jersey, limiting the applicability to other geographic regions, although the state’s population is diverse, and the network included 13-hospitals (both academic and community) all with differing COVID-19 treatment protocols. Lastly, we acknowledge the possibility of sampling bias as we collected data from a convenience sample in attempts to conduct the investigation quickly during this pandemic.

## Conclusions

This observational cohort study from a multi-hospital system spanning New Jersey, when taken together with other observational studies in the New York region, does not support the routine (off label and outside of clinical trial) use of hydroxychloroquine to reduce mortality among hospitalized COVID-19 patients. We cannot comment about the efficacy of this agent in the prehospital setting. Our exploratory review of IL-6 blockade with tocilizumab among ICU patients is encouraging and warrants further study. Common concerns regarding the limitations of observational studies should be applied until results of randomized trials are available.

## Data Availability

De-identified data is not currently available on a public repository.

## Acknowledgments

The authors wish to thank the nurses, data managers, and physicians who after caring for their patients assisted in the abstraction of the clinical data.

